# Community pharmacist-led medicines use review for asthma and COPD: a randomised controlled trial of effectiveness and cost-effectiveness

**DOI:** 10.1101/2025.03.12.25323845

**Authors:** Ethan Phillips, Lydia Prieto Sepulveda, Nunzio Crimi, Andrea Manfrin, Apostolos Tsiachristas

**Author notes:** Joint senior authorship (Andrea Manfrin, Apostolos Tsiachristas). Correspondence to A Manfrin.

## Abstract

**Background:** Prior studies have demonstrated the potential effectiveness of pharmacists assisting with chronic disease management for certain conditions, yet existing evidence is limited.

**Aim:** Evaluate the effectiveness and cost-effectiveness of a community pharmacist-led intervention to improve disease control for asthma and COPD patients.

**Design and Setting:** A pragmatic, two-arm, parallel randomised controlled trial (ISRCTN 38734433) across 100 community pharmacies in Sicily, Italy.

**Method:** Adult patients with asthma or COPD taking prescribed medications were randomised (2:1) to receive either pharmacist-led medicines use review consultations at baseline and six months (intervention) or usual care (control). Disease control was assessed at three-month intervals for a year using the Asthma Control Test (ACT≥20) or the Clinical COPD Questionnaire (CCQ<2). Health utility was estimated using the EQ-5D-5L. Missing non-baseline data were multiply imputed.

**Results:** 835 patients provided baseline data. Disease control increased by 6.3 percentage points in the intervention group compared with a 1.4 percentage-point decrease in the control group. The adjusted odds ratio for disease control at 12 months was 1.41 (95% CI 1.01 to 1.98; p=0.044). The intervention had an incremental cost of –€116.75 (95% CI –€1,366.77 to €1,133.27) and an incremental effect of 0.022 QALYs (95% CI –0.020 to 0.065) per patient over one year, with an 86.1% probability of being cost-effective at a willingness-to-pay threshold of €29,000 per QALY.

**Conclusion:** The pharmacist-led intervention was effective and likely to be cost-effective at improving disease control, supporting expanded utilisation of community pharmacists in chronic respiratory condition management.

**How it Fits In:** Prior studies suggest that community pharmacists can support chronic respiratory disease management, yet existing evidence is limited by short follow-up, insufficient economic evaluation, and single-disease samples. This pragmatic trial found that two pharmacist-delivered medicines use reviews improved disease control for adult asthma and COPD patients over 12 months and were likely to be cost-effective. Our results support greater involvement of community pharmacists in chronic disease management as part of multidisciplinary, neighbourhood-based primary health care services, while recognising that implementation may require adaptation to local healthcare contexts.

**Summary of Research:** Community pharmacist-led medicines use reviews improved 12-month asthma and COPD control and were likely cost-effective within the Italian healthcare system.

## INTRODUCTION

Chronic respiratory conditions (CRCs), including chronic obstructive pulmonary disease (COPD) and asthma, impose a substantial and growing burden on European health systems (1–6). Ageing populations and rising chronic disease prevalence are increasing healthcare utilisation and expenditure at a time when many European countries face primary care workforce shortages and constrained health budgets (3–7). Effective long-term management of CRCs requires regular monitoring, medication optimisation, and patient counselling to promote disease control (3,8,9). Multidisciplinary team-based models that redistribute appropriate activities to multiple healthcare professionals beyond general practice physicians may therefore be needed to expand service capacity and shift toward proactive, community-based CRC management (8,9).

Community pharmacists are accessible, neighbourhood-embedded healthcare professionals with the necessary pharmaceutical expertise to support medicine optimisation, adherence monitoring, patient education and counselling, and early identification of deterioration (10–12). However, they remain underutilised in primary health care service delivery (13–16). Primarily viewed as only dispensers rather than care providers, the contribution of community pharmacists to chronic disease management is limited by regulatory and licensing constraints, poor integration within primary care teams, high workloads, and misaligned remuneration frameworks (17).

Amidst increasing pressures on healthcare systems in Europe, evidence of value-for-money is critical for commissioning decisions (18). Although prior economic evaluations have found that pharmacist-delivered disease management and medication review services are cost-effective (19), evidence specific to asthma and COPD remains limited. Nevertheless, the UK’s National Institute for Health and Care Excellence (NICE) has suggested that community pharmacies can cost-effectively promote more accessible primary healthcare (20). This guidance is reflected in the UK’s “Pharmacy First” scheme which allows pharmacists to independently assess and treat seven common acute conditions (21). Elsewhere in Europe, similar programmes have allowed community pharmacists to take on greater roles within primary healthcare delivery, including disease screening, vaccination, health promotion, medications and adherence review, chronic condition management, first-time dispensing, and patient advising (17). However, further utilisation of community pharmacists in CRC management may still be hindered by a lack of reliable evidence, especially relating to the health outcomes and cost-effectiveness of expanded community pharmacist services (17).

Previous studies in Europe suggest that pharmacist-led interventions may improve medication adherence, inhaler technique, quality of life, or disease management in people with asthma or COPD (22–26). However, the existing evidence is limited by small sample sizes, short follow-up, and heterogeneous outcome measures. Many studies have only used process or activity measures rather than clinically relevant health outcomes. Furthermore, studies to date have rarely included comprehensive cost-effectiveness analyses using reliable healthcare cost data (22,24–26).

The earlier Italian I-MUR trial informed national investment in pharmacist-led services for adult asthma patients (26,27), suggesting that more robust evidence may be useful in expanding access to CRC management services for non-asthma patients and in other European countries. This study addresses limitations of previous research by evaluating the effectiveness and cost-effectiveness of a pharmacist-led medicines use review intervention to improve disease control among both asthma and COPD patients, collecting health outcomes over a longer 12-month study period, and using prospectively collected health system cost data for cost-effectiveness analysis.

## METHODS

We conducted a pragmatic, two-arm, parallel, randomised controlled trial in 100 community pharmacies in Sicily, Italy, following the published protocol and analysis plans (28). Consistent with the PRECIS-2 framework for pragmatic trials, the intervention was delivered in routine pharmacy settings, compared to usual care, enrolled patients through their normal patterns of healthcare engagement, and captured patient-centred outcomes over 12 months (29). Reporting follows the CONSORT 2025 guidelines for clinical trials and the CHEERS 2022 guidelines for health economic evaluations (30,31). Checklists are provided in the Supplementary Information.

### Study population and recruitment process

Pharmacies and pharmacists were screened according to the full eligibility criteria reported in the protocol (28). Participating pharmacies, mostly privately owned, were required to have a private consultation area and a telephone or other device for remote consultations (28). Pharmacies could not be involved in any other clinical research at the time of recruitment (28). Pharmacists were eligible if they were qualified and registered with the Italian Pharmacy Board, had experience advising patients and conducting disease monitoring activities, and attended a training session (28). Pharmacists provided informed consent to participate before patient recruitment began.

Adults with either asthma or COPD were recruited via convenience sampling through referral by recruiting pharmacies, primary care providers, or specialist physicians between May 2022 and June 2022. A recruitment target of 900 was set based on prospective power calculations reported in the protocol (28); however, the study was not powered to detect effects among disease subgroups (i.e. asthma alone or COPD alone) since the primary outcome was not condition-specific. Due to the convenience sampling method involving recruitment through routine care pathways, providers did not record the total number of patients assessed. Eligible participants were at least 18 years old, had been diagnosed with asthma or COPD for at least six months, and were taking prescribed medication for their condition (28). Patients were excluded if they had a terminal illness (either reported or identified via prescription coding), were enrolled in another clinical trial, relied on another person to administer their medications, or were unable to communicate well in written and spoken Italian (28). After eligibility was assessed, patients were given informational materials and up to a week to consider participation. Those willing to participate provided written informed consent.

At the time of recruitment, participants were individually randomised on a 2:1 basis to either receive the intervention or usual care (28). Individual rather than cluster randomisation was selected to preserve statistical efficiency and feasibility during pandemic-era recruitment. Although the earlier I-MUR trial suggested benefit in asthma (26,27), uncertainty remained because CRC-MUR differed in its broader target population, consultation schedule, and follow-up, thus preserving clinical equipoise for the adapted intervention. The 2:1 allocation ratio increased the proportion receiving CRC-MUR while retaining a usual-care comparison. The sample size calculations accounted for unequal allocation but not pharmacy-level clustering

(28). A computer-randomised patient ID was assigned to each participant automatically using a pre-set list of possible IDs in the central FarmPro Care platform. Each ID had a corresponding pre-randomised group assignment and pharmacists could not view the next allocation to avoid any manipulation (unconscious or otherwise) of the recruitment process. Neither patients nor pharmacists delivering the consultations could be blinded, but investigators and study administrators remained blinded to allocation.

### Intervention

Patients assigned to the intervention group received the Chronic Respiratory Condition Medicines Use Review (CRC-MUR) intervention. CRC-MUR was adapted from the asthma-specific Italian Medicines Use Review (I-MUR) for use among adults with asthma or COPD (26–28). CRC-MUR comprised structured one-to-one consultations with community pharmacists at baseline (T0) and six months (T6) to review symptoms, health and social care utilisation, medication use, medication acceptance and adherence, and any pharmaceutical care issues (28). Pharmacists provided condition and lifestyle counselling and communicated relevant findings or recommendations to patients’ general practitioners or specialists to support continuity of care and minimise the risk of inconsistent clinical advice. In most cases, patients received the intervention from the same pharmacist in both instances, unless unavailable due to illness. Consultations were conducted in-person when possible, but a small number were delivered remotely via phone or video call due to COVID-19 restrictions. The mode of delivery for individual consultations was not systematically recorded and therefore could not be incorporated into the effectiveness or cost-effectiveness analyses. Participating pharmacists received standardised training on procedures for data collection, intervention delivery, patient advising, and follow-up communication with patients’ usual care providers. This training helped to improve intervention quality and uniformity across participating sites and pharmacists and reduced potential observer bias.

Participants assigned to the control group received usual care without structured CRC-MUR counselling and advice. Allocation to the control group did not alter participant medications or care provision and did not change pharmacists’ professional responsibilities. Pharmacists continued to respond to clinically important concerns according to normal practice, including referral to the patient’s usual care provider where appropriate. Pharmacists were instructed to separate the times they interacted with patients from different study arms to reduce the potential for contamination. Pharmacists were also provided with arm-specific Gantt charts to ensure compliance with the correct schedule of intervention delivery and data collection.

Implementation of CRC-MUR required trained community pharmacists, dedicated consultation time, access to private consultation space, and mechanisms for communicating with other healthcare providers. The economic evaluation included the cost of pharmacist consultation time, while training, facilities, and routine communication infrastructure were considered part of existing service capacity and were not separately costed.

### Data Collection

Patients completed a survey at baseline (T0) to collect demographic data and patient characteristics. Pharmacists recorded outcome data using a standardised questionnaire administered to participants in both treatment groups at baseline visit (T0) and three-month intervals for the following year (T3, T6, T9, and T12). Intervention participants received consultation and advice alongside the questionnaire, while control participants received no feedback or guidance.

Health outcome data were collected using the Asthma Control Test (ACT) for asthma patients, ranging from 5 to 25 with higher scores indicating better disease control, or Clinical COPD Questionnaire (CCQ) for COPD patients, ranging from 0 to 6 with lower scores indicating better disease control (32,33). The EQ-5D-5L questionnaire was used to capture patient quality-of-life (34).

After each visit, pharmacists entered collected data into the secure FarmPro Care platform. All electronic patient data were stored using anonymised study IDs in accordance with applicable data protection requirements. Information linking participants to their study IDs was available only to the pharmacists. Signed informed consent forms were stored securely at the patient’s recruiting pharmacy.

This study used a health system cost perspective with a 12-month cost horizon. Monthly cost data in Euros were obtained for each patient from the Sicilian Regional Health Service Database from June 2022 to June 2023 and categorised as outpatient care, inpatient care, laboratory tests, medication, or medication delivery. These data included all costs incurred by the Italian national health service but did not capture out-of-pocket payments or payments made by private health insurers. No discounting was applied since all costs were incurred in the same year.

### Patient and Public Involvement

Direct engagement with patients during study design was not feasible due to the COVID-19 pandemic. However, the Italian patient advocacy organisation CittadinanzAttiva (Active Citizenship) reviewed the research protocol for clarity, participant burden, and practical relevance. Their review led to an additional questionnaire item asking whether participants lived alone, reflecting concerns about social isolation and support during the pandemic. Overall, the organisation gave positive feedback regarding the proposed research.

### Outcomes

The primary outcome was disease control at 12 months, defined prospectively as ACT ≥20 for asthma patients or CCQ <2 for COPD patients (28,32,33). This binary definition provided a clinically interpretable common outcome across the two conditions, although it discarded some information from the continuous scores. Secondary outcomes were disease control at intermediate time points (3, 6, and 9 months) and cost-effectiveness assessed using health system costs and quality-adjusted life years (QALYs) per patient over the study period.

### Missing Data Analysis & Imputation

To interrogate the missing-at-random assumption, we examined whether completeness of effectiveness and cost-effectiveness data was associated with treatment allocation, demographic characteristics, diagnosis, baseline disease control, or quality of life. Completeness for effectiveness data was defined as having no missing data for sex, age, quality-of-life at baseline, and ACT or CCQ scores at all time points. Completeness for cost-effectiveness data additionally required no missing data for 6– and 12-month costs across all categories.

We employed multivariate imputation by chained equations (MICE) to fill missing post-baseline disease control, quality-of-life scores, and cost data. Twenty imputed datasets were generated, exceeding the proportion of missing observations according to MICE best practice (35). Predictions were based on demographic characteristics, baseline health, previous disease-control and quality-of-life scores, healthcare utilisation, and costs where relevant. Imputation was performed separately within treatment arms to avoid treatment-effect leakage, and future observations were not used to impute earlier outcomes. Post-hoc sensitivity analyses using complete-cases, simple imputation, and alternative MICE predictor matrices examined sensitivity to missing-data assumptions and imputation methods. Full details are provided in the Supplementary Information.

### Health Outcome Analysis

Following the prespecified intention-to-treat (ITT) approach, we used logistic generalised estimating equations (GEE) to assess the effect of the intervention on disease control at 12 months (primary outcome) and intermediate time points (secondary outcomes). Results from each of the 20 imputed datasets were pooled according to Rubin’s rules (36). The regression was run first unadjusted and then adjusted for sex, age, and disease control at baseline. Post-hoc robustness checks and sensitivity analyses explored alternative covariate adjustment specifications, cross-sectional logistic regression, and mixed-effects models with patient-level and pharmacy-clustered random intercepts. Further details are provided in the Supplementary Information.

### Cost-Effectiveness Analysis

Using the multiply-imputed data to follow the ITT approach, 12-month health system costs were summed across categories. Intervention costs of €80 (€40 per consultation, in 2022 euros) were added for intervention participants to account for the community pharmacist time spent delivering the intervention, derived from the earlier I-MUR study costing framework. Post-hoc sensitivity analyses evaluating higher intervention costs and alternative approaches to handling missing cost data are described in the Supplementary Information.

EQ-5D-5L responses were valued using the Italian tariff set and summed across domains to calculate quality-of-life index scores ranging from 0 (health state equivalent to death) to 1 (complete health) at each study time point (37). A post-hoc sensitivity analysis using the United Kingdom value set is presented in the Supplementary Information (38). QALYs were estimated using trapezoidal integration of quality-of-life (utility) over the study period.

Incremental costs and QALYs were estimated using Gaussian identity-link generalised linear models adjusted for sex and age. Cost-effectiveness was evaluated across willingness-to-pay thresholds and reported at €29,000 per QALY, approximately equivalent to £25,000 per QALY. Economic uncertainty was assessed using 5,000 treatment-stratified bootstrap samples. Results for each bootstrap sample were pooled across imputed datasets to generate incremental cost and QALY estimates for each iteration. Resulting iteration-level estimates were used to construct the cost-effectiveness plane and cost-effectiveness acceptability curve.

### Data Analysis Software

All data transformations and analyses were conducted in R version 4.4.1, using the tidyverse (v2.0.0), dplyr (v1.1.4), geepack (v1.3.13), lme4 (v1.1-35.5), and mice (v3.16.0) packages (39–47).

## RESULTS

### Patient Characteristics

Between May 2022 and June 2022, 836 participants were enrolled. Final data collection was completed in June 2023. One participant lacked baseline data, resulting in 835 participants eligible for analysis. Of those, 409 (49.0%) patients had asthma and 426 (51.0%) had COPD. Among asthma patients, 266 (65.0%) received the intervention and 143 (35.0%) received usual care; among COPD patients, 294 (69.0%) received the treatment and 132 (31.0%) received usual care. Figure 1 shows the CONSORT flow diagram.

**Figure 1.**
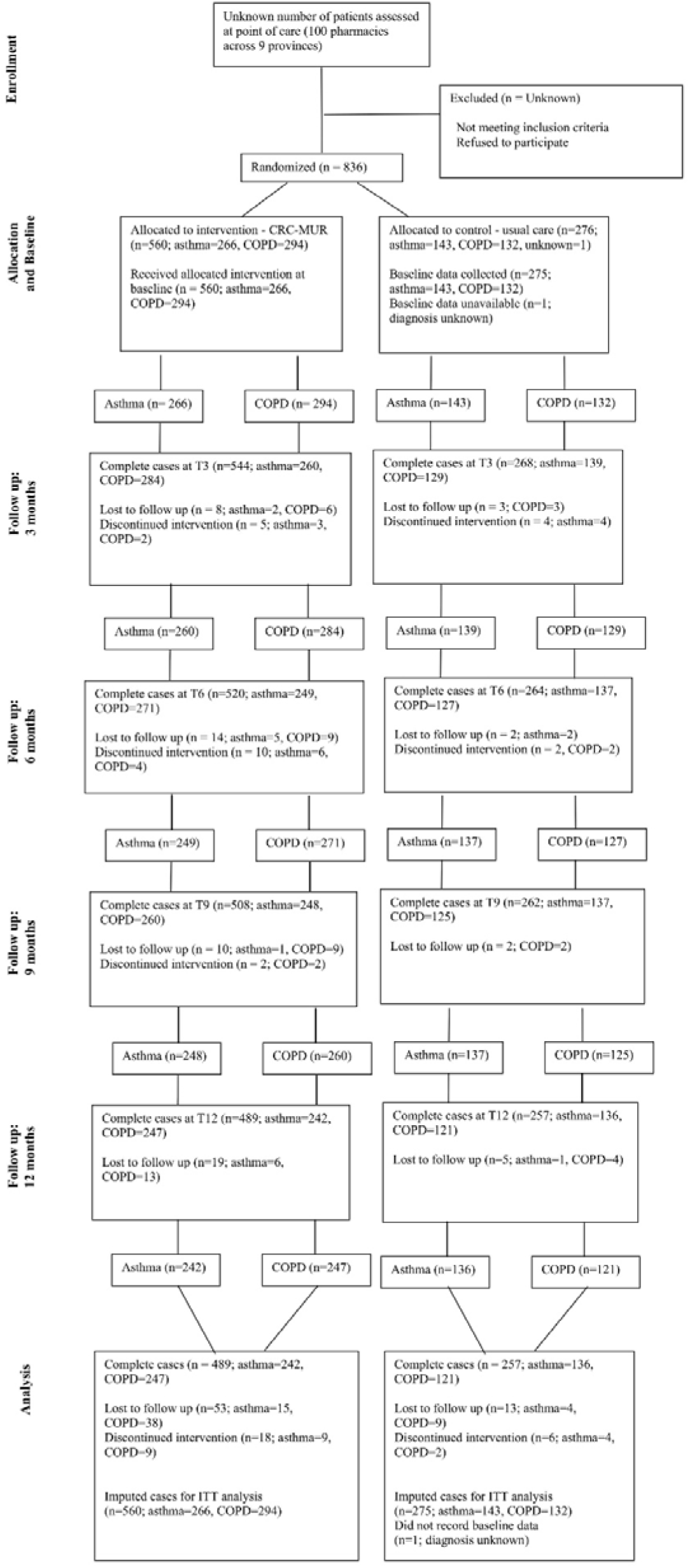
CONSORT flow diagram of participant recruitment, allocation, and follow-up.

The treatment groups were broadly comparable across demographic characteristics at baseline; standardised differences were below 0.10 for all but one characteristic. Baseline differences in disease severity between condition subgroups are consistent with chance variation and were addressed by model adjustment for baseline disease control. Table 1 shows the baseline demographic and condition characteristics of study participants by treatment arm. More detailed characteristics by condition are reported in Supplementary Table 1.

**Table 1.**
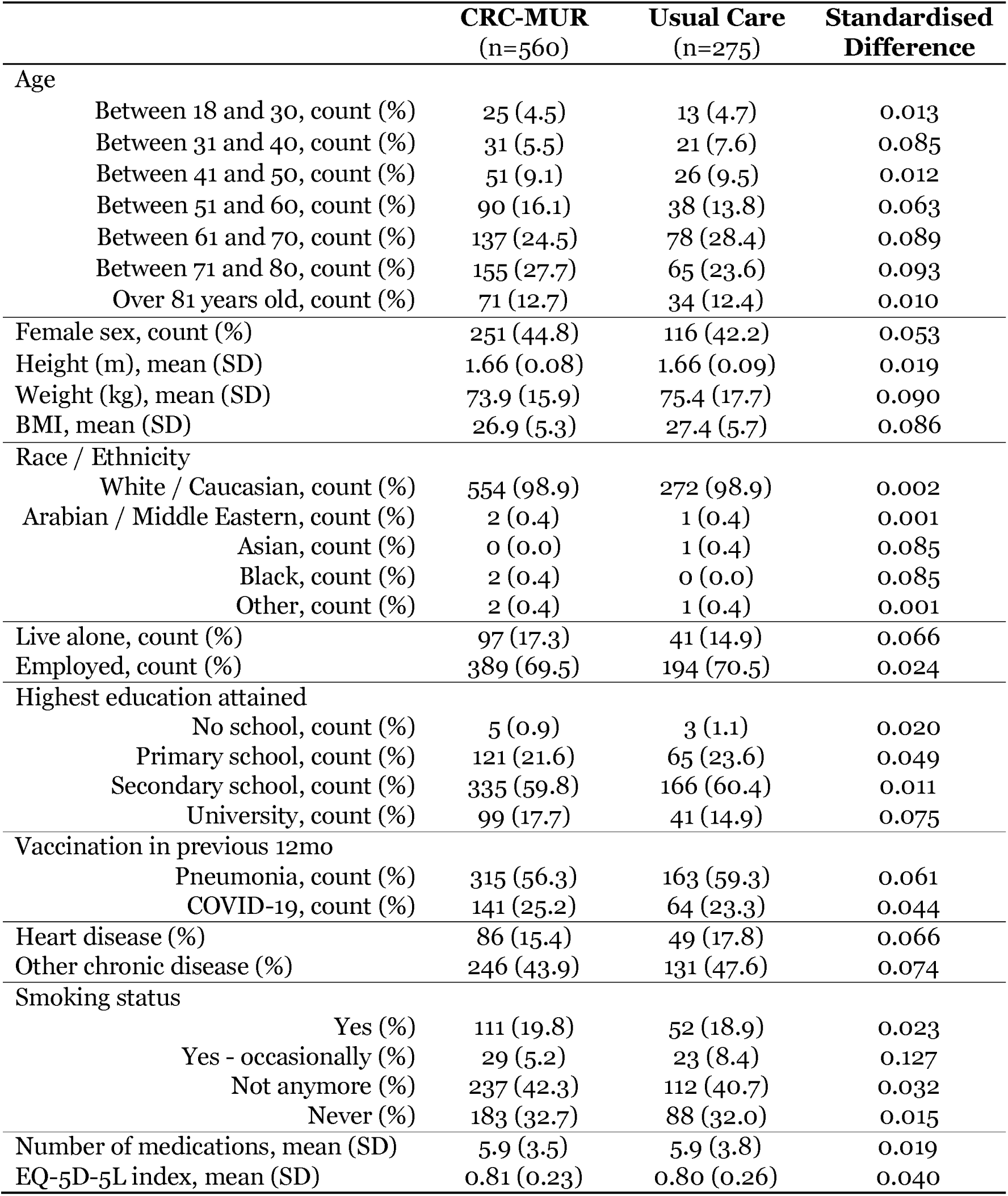
Baseline characteristics of patients by treatment arm.

|  | **CRC-MUR**<br>(n=560) | **Usual Care**<br>(n=275) | **Standardised**<br>**Difference** |
| --- | --- | --- | --- |
| **Age** |  |  |  |
| Between 18 and 30, count (%) | 25 (4.5) | 13 (4.7) | 0.013 |
| Between 31 and 40, count (%) | 31 (5.5) | 21 (7.6) | 0.085 |
| Between 41 and 50, count (%) | 51 (9.1) | 26 (9.5) | 0.012 |
| Between 51 and 60, count (%) | 90 (16.1) | 38 (13.8) | 0.063 |
| Between 61 and 70, count (%) | 137 (24.5) | 78 (28.4) | 0.089 |
| Between 71 and 80, count (%) | 155 (27.7) | 65 (23.6) | 0.093 |
| Over 81 years old, count (%) | 71 (12.7) | 34 (12.4) | 0.010 |
| Female sex, count (%) | 251 (44.8) | 116 (42.2) | 0.053 |
| Height (m), mean (SD) | 1.66 (0.08) | 1.66 (0.09) | 0.019 |
| Weight (kg), mean (SD) | 73.9 (15.9) | 75.4 (17.7) | 0.090 |
| BMI, mean (SD) | 26.9 (5.3) | 27.4 (5.7) | 0.086 |
| **Race / Ethnicity** |  |  |  |
| White / Caucasian, count (%) | 554 (98.9) | 272 (98.9) | 0.002 |
| Arabian / Middle Eastern, count (%) | 2 (0.4) | 1 (0.4) | 0.001 |
| Asian, count (%) | 0 (0.0) | 1 (0.4) | 0.085 |
| Black, count (%) | 2 (0.4) | 0 (0.0) | 0.085 |
| Other, count (%) | 2 (0.4) | 1 (0.4) | 0.001 |
| Live alone, count (%) | 97 (17.3) | 41 (14.9) | 0.066 |
| Employed, count (%) | 389 (69.5) | 194 (70.5) | 0.024 |
| **Highest education attained** |  |  |  |
| No school, count (%) | 5 (0.9) | 3 (1.1) | 0.020 |
| Primary school, count (%) | 121 (21.6) | 65 (23.6) | 0.049 |
| Secondary school, count (%) | 335 (59.8) | 166 (60.4) | 0.011 |
| University, count (%) | 99 (17.7) | 41 (14.9) | 0.075 |
| **Vaccination in previous 12mo** |  |  |  |
| Pneumonia, count (%) | 315 (56.3) | 163 (59.3) | 0.061 |
| COVID-19, count (%) | 141 (25.2) | 64 (23.3) | 0.044 |
| Heart disease (%) | 86 (15.4) | 49 (17.8) | 0.066 |
| Other chronic disease (%) | 246 (43.9) | 131 (47.6) | 0.074 |
| **Smoking status** |  |  |  |
| Yes (%) | 111 (19.8) | 52 (18.9) | 0.023 |
| Yes - occasionally (%) | 29 (5.2) | 23 (8.4) | 0.127 |
| Not anymore (%) | 237 (42.3) | 112 (40.7) | 0.032 |
| Never (%) | 183 (32.7) | 88 (32.0) | 0.015 |
| Number of medications, mean (SD) | 5.9 (3.5) | 5.9 (3.8) | 0.019 |
| EQ-5D-5L index, mean (SD) | 0.81 (0.23) | 0.80 (0.26) | 0.040 |

### Missingness Analysis

Before imputation, effectiveness data were complete for 746 patients (89.3%), and economic data were complete for 802 patients (96.0%). Effectiveness data completeness was weakly but statistically significantly associated with treatment allocation and baseline quality of life. There were no statistically significant predictors of economic data completeness.

### Intervention Effectiveness

Disease control increased by 6.3 percentage points (95% CI [2.2, 10.4]) in the CRC-MUR group and decreased by 1.4 percentage points (95% CI [-7.3, 4.5]) in the control group between baseline and 12 months. In relative terms, this represents a 12.3% increase in disease control among intervention recipients, compared to a 2.6% decrease for usual care. Table 2 shows the proportion of patients with successful disease control at each time point, as well as change in disease control across the study period. Outcomes by condition, including average CCQ, ACT, and EQ-5D-5L scores, are reported in Supplementary Tables 2 and 3. Average changes in ACT and CCQ scores were observed to be below the minimal clinically important differences (48,49).

**Table 2.** Proportion of patients with successful disease control across study time points.

|  | **CRC-MUR**<br>(n = 560) | **Usual Care**<br>(n = 275) |
| --- | --- | --- |
| Baseline, % (SE) | 51.1 (2.1) | 53.5 (3.0) |
| 3-months, % (SE) | 56.4 (2.1) | 50.1 (3.0) |
| 6-months, % (SE) | 56.8 (2.1) | 51.7 (3.0) |
| 9-months, % (SE) | 56.0 (2.1) | 49.6 (3.0) |
| 12-months, % (SE) | 57.4 (2.1) | 52.1 (3.0) |
| Change from baseline to 12 months, percentage points (SE) | 6.3 (2.1)* | -1.4 (3.0) |

\*Significant ( $p < 0.05$ )
Note: Significance was tested for within-group change from baseline to 12 months.

**Table 3.**
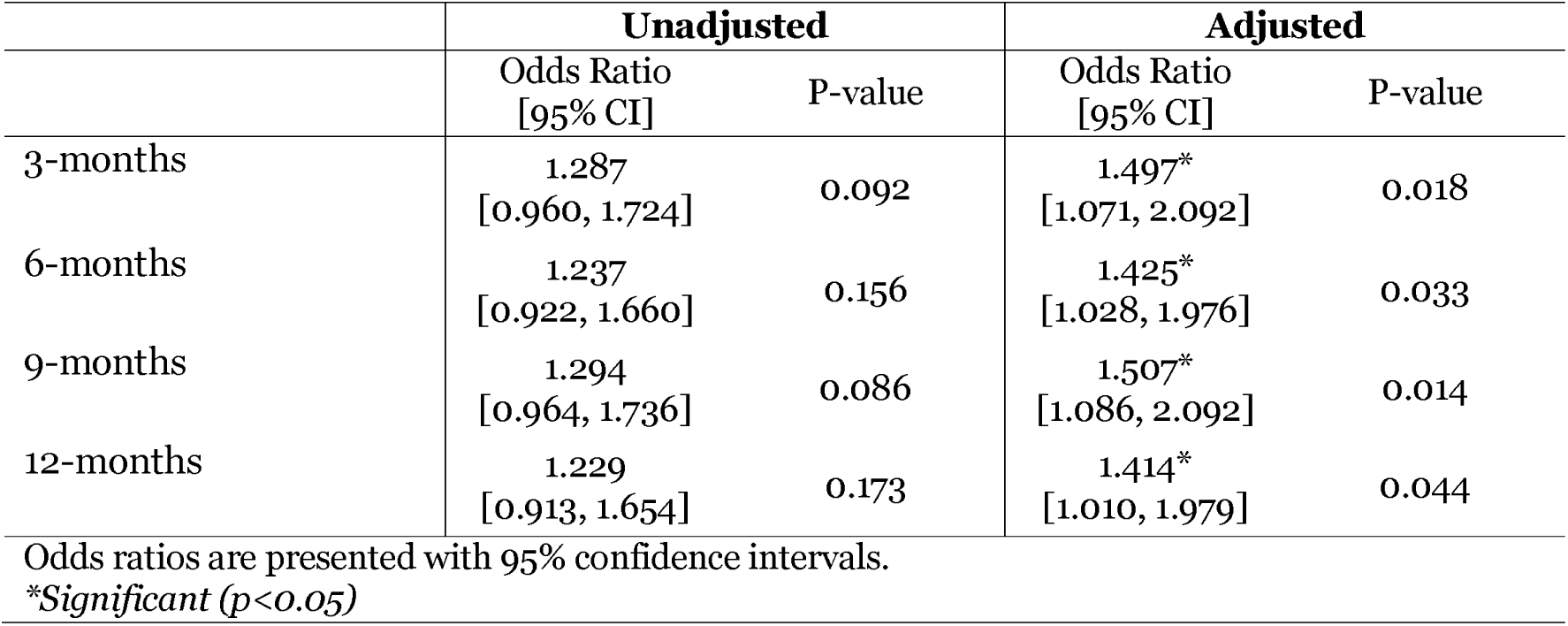
GEE regression results for effect of CRC-MUR on odds of disease control across study time points.

The adjusted odds ratio for disease control at 12 months was 1.41 (95% CI [1.01, 1.98]; p = 0.044) for CRC-MUR compared to usual care, adjusting for sex, age, and disease control at baseline. Figure 2 displays the adjusted odds ratios for disease control over the study period, and Table 3 presents both the unadjusted and adjusted odds ratios at each time point.

**Figure 2.**
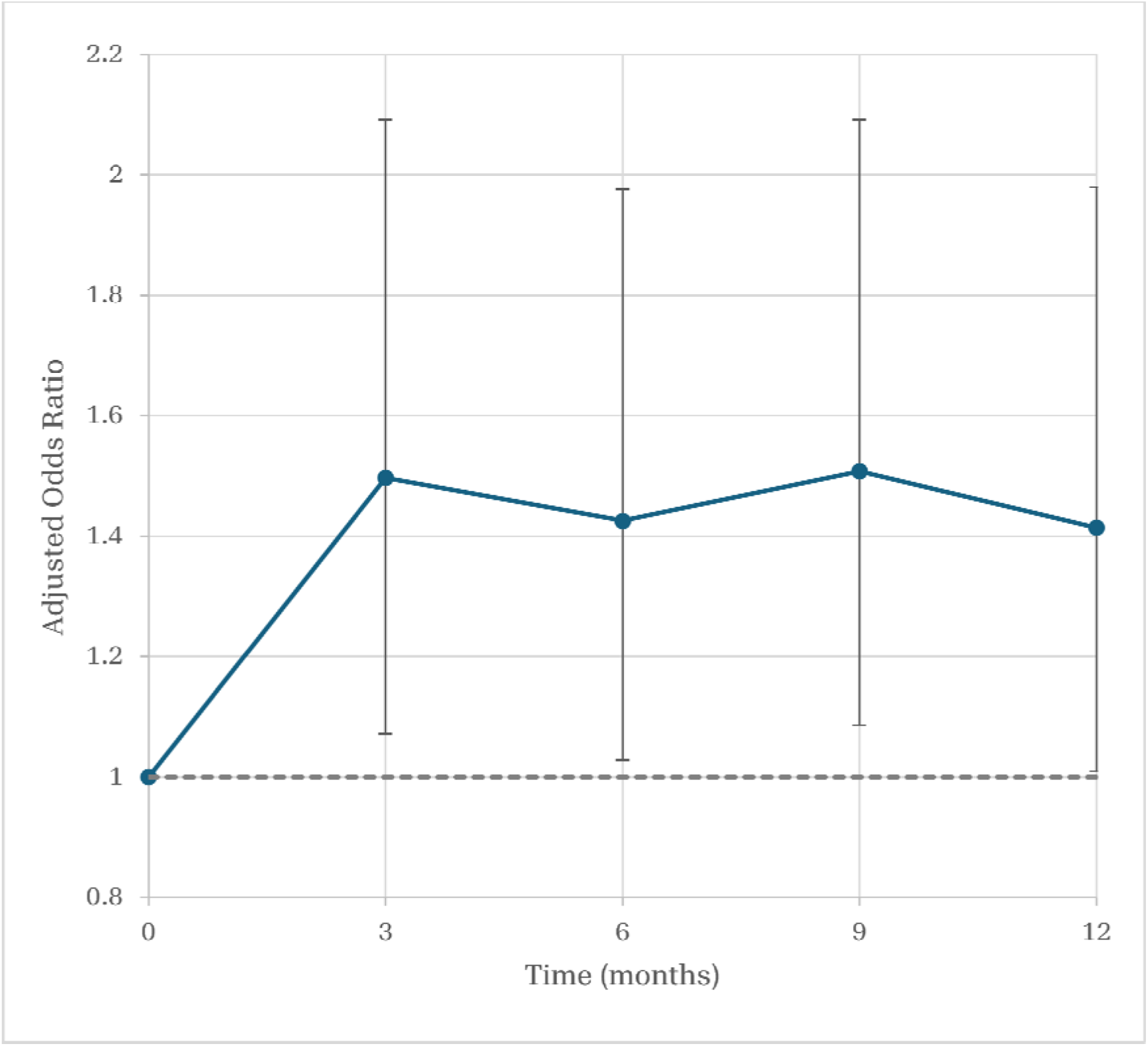
Adjusted odds ratios for disease control for CRC-MUR versus usual care over 12 months. Estimates favoured CRC-MUR and were statistically significant at all follow-up time points; error bars show 95% confidence intervals.

### Resource Utilisation

Although the intervention group did report marginally lower average healthcare utilisation across most categories over the year and lower average total costs compared to the control group, none of the between-group differences were statistically significant. The intervention group reported more therapist visits in the six months prior to baseline, but this difference did not persist. Detailed health and social care utilisation results are presented in Supplementary Table 4. Health system costs by category are compared across study arms in Supplementary Table 5.

### Cost-Effectiveness Analysis

CRC-MUR was associated with an incremental cost of –€117 (95% CI [-€1,370, €1,130]) and an incremental effect of 0.022 QALYs (95% CI [-0.020, 0.065]). Although neither estimate was statistically significant, these results indicate that the intervention resulted in lower average costs and higher average health-related quality of life than usual care. The probability of cost-effectiveness was 86.1% at a threshold of €29,000 per QALY (approximately equal to £25,000 per QALY).

Figure 3 presents the cost-effectiveness plane, with the pooled point estimate in the southeast quadrant. Supplementary Figure 1 shows the acceptability curve across a range of willingness-to-pay thresholds.

**Figure 3.**
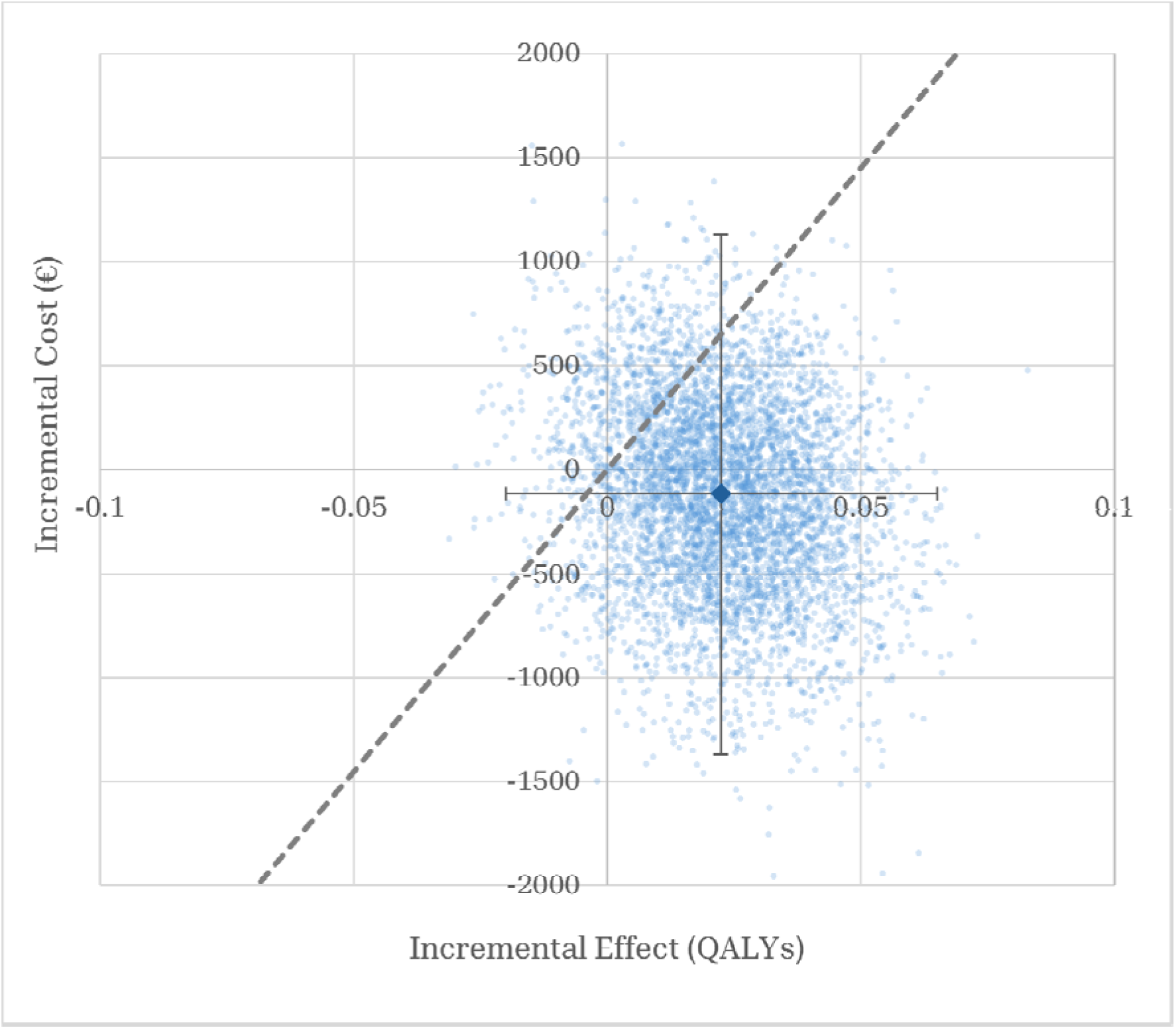
Cost-effectiveness plane for CRC-MUR. Each small dot represents the result pooled across 20 imputed datasets for one bootstrap iteration; the larger diamond represents the mean across bootstrap iterations. The dashed line represents a cost-effectiveness threshold of €29,000 per QALY (approximately £25,000 per QALY).

### Robustness and Sensitivity Analyses

Effectiveness findings were directionally consistent and favourable to CRC-MUR across alternative approaches for missing data, covariate specifications, regression models, and clustering. The estimated pharmacy-level latent scale intraclass correlation coefficient (ICC) was 0.132 (range across imputations 0.119–0.143), but incorporating pharmacy-level random effects yielded findings that were highly consistent with the primary analysis.

The economic conclusions also remained consistent across alternative missing data approaches, EQ-5D-5L tariff sets, and intervention cost assumptions. The probability of cost-effectiveness remained above 70% in all intervention cost scenarios.

Detailed results for all sensitivity analyses are presented in Supplementary Tables 6-10.

## DISCUSSION

### Summary

We found that the pharmacist-led intervention, CRC-MUR, had a statistically significant, positive effect on improving disease control as early as three months after randomisation, and this effect persisted throughout the 12-month study period. Despite small variations in the effect size across follow-up time points, the intervention effect favoured CRC-MUR at all time points in the study period. However, confidence intervals for adjusted effectiveness estimates were wide and the lower bound at 12 months was close to the null. Therefore, the magnitude and precision of the intervention’s effectiveness should be interpreted cautiously.

Although the absolute improvement in disease control was modest (approximately six percentage points over 12 months), it was achieved using only two pharmacist consultations during the study period. Given the low-intensity nature of the intervention and the large number of patients with asthma and COPD managed in primary care, even modest improvements may translate into meaningful population-level health gains if implemented at scale. However, mean changes in the continuous ACT and CCQ scores remained below their established minimal clinically important differences, so the statistical effect on the binary disease-control outcome should not be interpreted as a large average improvement in symptom scores.

The CRC-MUR intervention was likely to be cost-effective, with an 86.1% probability of cost-effectiveness at a willingness-to-pay threshold of €29,000 per QALY. Point estimates indicated improved quality of life (approximately eight additional quality-adjusted days over one year) and lower healthcare costs compared to usual care. However, the confidence intervals for both incremental costs and QALYs included the null, so economic findings should be interpreted as favourable but uncertain.

Importantly, these findings were observed within a pragmatic trial conducted under routine community pharmacy conditions, supporting the applicability of the intervention to real-world primary care settings. The effectiveness and cost-effectiveness were also robust to alternative modelling, imputation, clustering, and economic assumptions evaluated through sensitivity analyses.

### Strengths and Limitations

A major strength of this study is its pragmatic design, intended to maximise applicability to routine community pharmacy practice. However, several of these same pragmatic design choices reduced control over participant selection and intervention delivery, limiting sample representativeness and generalisability.

A further limitation is that neither patients nor pharmacists could be blinded to treatment allocation due to the nature of the intervention. This may have introduced performance bias, whereby participants receiving CRC-MUR became more engaged with management of their condition or more likely to report favourable outcomes. Because disease control, healthcare utilisation, and quality of life were patient-reported outcomes, knowledge of treatment allocation may also have influenced recall or reporting. However, ACT and CCQ are both validated, clinically relevant measures of CRD patient health outcomes and the EQ-5D-5L is a robust, validated measure of health-related quality of life. Nevertheless, allocation-related reporting bias may have affected both the effectiveness findings and incremental QALY estimate in the economic evaluation.

Repeated participant contact throughout the study may have increased awareness of health management and encouraged behavioural changes independent of the intervention itself, although many follow-up assessments coincided with routine pharmacy visits for medication collection and therefore did not require substantial additional healthcare interactions beyond usual care. Additionally, follow-up procedures were the same for both treatment arms.

Because the same pharmacists managed participants in both groups, intervention practices may have influenced usual-care interactions. Training and arm-specific schedules were used to reduce contamination, but intervention fidelity was not formally monitored. If present, such contamination would be expected to bias estimates towards the null.

Since patients recruited through the same pharmacy may share pharmacists, local healthcare environments, and referral patterns, residual pharmacy-level correlation was possible despite individual randomisation. Sensitivity models incorporating pharmacy-level random effects identified modest pharmacy clustering but produced treatment estimates consistent with the primary GEE analysis, suggesting that within-pharmacy correlation was unlikely to have meaningfully influenced the study conclusions. The trial was also not powered to detect condition-specific treatment effects. Intervention effectiveness for asthma or COPD patients alone should therefore be interpreted cautiously, and possible heterogeneity between conditions requires further investigation.

This study adopted a health system costing perspective and therefore excluded out-of-pocket expenditure, informal care, productivity losses, and privately funded healthcare. Consequently, the economic evaluation may underestimate the total societal costs. Additionally, the 12-month study period means that long-term clinical effects and cost-effectiveness remain uncertain beyond one year.

The supportive involvement of the Sicilian region enabled us to collect reliable cost data directly from the health system database. However, the relative demographic homogeneity of the Sicilian study population may limit generalisability. Convenience recruitment through routine care pathways also limits the external validity of our findings. Patients with more frequent pharmacy or healthcare provider encounters may have had more opportunities to be recruited, while individuals with severe disability, reliance on carers, or work and family commitments may have been underrepresented. Transferability will also depend on primary healthcare system structure, pharmacist scope of practice, reimbursement mechanisms, and population characteristics. While the results of this study are likely generalisable to similar populations and healthcare systems elsewhere in Europe, further investigation with more diverse participant groups is needed.

Finally, this study was conducted during extremely challenging conditions brought on by the COVID-19 pandemic. Due to restrictions imposed during the pandemic, some pharmacist consultations had to be delivered via phone and video calls instead of the originally intended in-person format. Because the mode of delivery was not systematically recorded, we were unable to assess whether remote and in-person consultations differed in their effectiveness. Further research should evaluate whether mode of delivery influences implementation processes or patient outcomes.

### Comparison with Existing Literature

Our findings are consistent with existing literature on community pharmacist-led interventions for chronic disease management, while also providing more robust evidence than was previously available for utilising community pharmacists to assist with chronic disease management. Hudd (2020) highlighted pharmacists’ emerging role in managing patients with COPD, but unlike the results presented here, their analysis included neither disease control nor the economic impact of pharmacist activities (10). In a systematic review of pharmacist-led interventions for medication adherence and inhalation techniques for patients with asthma and COPD, researchers found that significant improvement in medication adherence was only observed in COPD patients (25). The same review suggested that more research was needed to establish a stronger evidence base for effective CRC interventions, especially among asthmatic patients (25); this trial contributes to the strength of the evidence base in this area, suggesting that MUR services lead to improved health outcomes and quality of life for both asthma and COPD patients.

Contrary to our findings, a cluster-randomised controlled trial of integrated disease management in primary care for COPD patients found no difference between the control and the intervention groups (50). However, the study in question did not involve community pharmacists in the intervention delivery, offering a potential explanation for the difference in observed effect size.

For asthma patients, pharmacist-led interventions have been evaluated in several countries, yet recent systematic reviews have not identified any UK studies on the effectiveness and cost-effectiveness of similar interventions (27,51–54). A 2019 scoping review identified 28 studies in the UK, including a pharmacist-led MUR intervention. However, none of them evaluated its effectiveness (55). The same review identified only one study supporting the cost-effectiveness of a pharmacist-led intervention: a new medicine service (NMS) with consultations for patients starting new medications. Unlike CRC-MUR, the NMS intervention evaluated was not specific to CRC patients and focused on patient adherence to prescribed medications, rather than long-term health outcomes (56). The patient follow-up for the NMS study was 10 weeks (56); however, a 26-week follow-up evaluation did not demonstrate effectiveness, consistency, or replicability (57).

The current study was informed by the chief investigator’s previous I-MUR study conducted in Italy, which confirmed the effectiveness and cost-effectiveness of pharmacist-led MUR for adult asthma patients (26). We have now demonstrated that pharmacists can effectively deliver the intervention to patients with different primary diagnoses. This current study further differs from the prior I-MUR trial in length (I-MUR: 9 months, CRC-MUR: 12 months), number of consultations (I-MUR: once at T0, CRC-MUR: twice at T0 and T6), and delivery format (I-MUR: in-person only, CRC-MUR: mix of in-person and remote due to COVID-19). The sustained positive effect indicates that additional pharmacist-led consultations beyond the first session may be useful, and their benefits are persistent over a full year.

This study extends previous evidence by demonstrating effectiveness in a combined asthma and COPD patient population over a longer follow-up period and under routine community pharmacy conditions. To our knowledge, this is also the first study to evaluate both the effectiveness and cost-effectiveness of a pharmacist-led medicines use review intervention across a combined population of patients with asthma and COPD using prospectively collected health-system cost data. This extends the economic evidence base beyond condition-specific studies and provides insight into the potential value of pharmacist-led chronic respiratory disease management across multiple conditions.

### Implications for Practice & Policy

CRC-MUR could allow community pharmacists to take on larger roles within multidisciplinary primary health care for patients with CRCs. At a health system-level, engaging these underutilised providers may support more efficient use of limited resources while improvements in disease control could theoretically reduce demand for acute healthcare services. However, we did not observe any significant difference in self-reported health care utilisation or health system costs between study arms within the first year of treatment. Therefore, longer follow-up will be needed to determine whether the observed benefits translate into sustained economic gains at scale.

The earlier asthma-specific I-MUR trial had major impacts on policy and practice through the Italian government’s nation-wide rollout of community pharmacist services (26,27). The observed effectiveness and likely cost-effectiveness of CRC-MUR warrant consideration of such interventions as part of broader strategies to improve chronic respiratory disease management across multiple conditions. This study also provides evidence supporting an expanded role for community pharmacists within multidisciplinary primary care teams, beyond their typical dispensing role. Structured medication review and patient counselling, in collaboration with existing primary care providers, may be transferable to other healthcare systems, including the UK.

However, implementation in other settings would depend on local factors such as regulatory frameworks, scope of practice for community pharmacists, workforce capacity, reimbursement arrangements, and integration with existing primary care services. Implementation costs, including pharmacist training, should be considered alongside the potential clinical and economic benefits when evaluating wider adoption of such services. Implementation should also consider how pharmacist-led services can better incorporate patient preferences and equity considerations, ensuring accessibility for underserved populations and avoiding exacerbation of existing health inequalities related to language, socioeconomic circumstances, or geographic location.

Further research should evaluate community pharmacist-led MUR interventions for other chronic conditions, examine implementation across different healthcare systems, and assess longer-term effects on healthcare utilisation and costs. Although the pharmacist-led intervention may have potential applicability beyond chronic respiratory disease, its effectiveness and cost-effectiveness are likely to vary according to disease complexity, medication requirements, consultation frequency, and the organisation of care.

## Declaration of Competing Interests

All authors have completed the ICMJE uniform disclosure form at http://www.icmje.org/disclosure-of-interest/ and declare: financial support for the study associated with this submitted work was provided by SOFAD srl. EJP and AT both declare ongoing research supported by pharmaceutical companies and NHS organisations in England and Wales, although these relationships are unrelated to this work and had no impact on their contribution to the present study. All other authors declare no financial relationships with any organisations that might have an interest in the submitted work in the previous three years and no other relationships or activities that could appear to have influenced the submitted work.

## Patient and Public Involvement

While restrictions relating to the COVID-19 pandemic made it impractical to engage directly with patients during the study design phase, we sought out and received feedback on the study protocol and aims from the Italian public advocacy group CittadinanzAttiva (Active Citizenship).

## Data and Code Availability

The patient data used in this study are available in an anonymised format on the Oxford University Research Archive (ORA). It can be accessed at the following DOI address: https://dx.doi.org/10.5287/ora-rjkzjgg4k.

As described in the data analysis software paragraph of the methods section, the code for this study was written in R version 4.4.1 using the tidyverse (v2.0.0), dplyr (v1.1.4), geepack (v1.3.13), lme4 (v1.1-35.5), and mice (v3.16.0) packages. All analysis code is available via GitHub at https://github.com/ethanp274/BOFE_trial.

## Use of Artificial Intelligence Tools

ChatGPT (OpenAI, versions 5.4-5.6) was used to assist with statistical programming support, code revision, manuscript editing, and responses to reviewer comments. AI-generated content was used only as a drafting and revision aid. All outputs were reviewed, fact-checked, and revised by the authors, who maintained full control over all scientific decisions and interpretations. The authors accept full responsibility for the integrity, accuracy, and originality of the work.

## Trial Registration

This study was prospectively registered as ISRCTN 38734433 (https://www.isrctn.com/ISRCTN38734433). The protocol was published at https://www.doi.org/10.1097/MD9.0000000000000158.

## Ethics Approval

Ethics approval for this study was obtained from relevant ethical review committees in both Italy and the United Kingdom.

Approved 22/02/2021, Azienda Ospedaliero-Universitaria Policlinico “G. Rodolico – San Marco” Catania (via Santa Sofia 78 – 95123 Catania, Italy; +39 (0)95 3781855) ref: 47/2021/PO

Approved 29/03/2021, University of Central Lancashire (Preston Fylde Road, PR1 2HE, UK; +44 (0)1772 895583) ref: HEALTH 0163

## Confirmation of Transparency

All authors affirm that the manuscript is an honest, accurate, and transparent account of the study being reported; that no important aspects of the study have been omitted; and that any discrepancies from the study as originally planned (and, if relevant, registered) have been explained.

## Role of Funding Source

This study was supported financially by SOFAD srl, a pharmacist-operated pharmaceutical distribution company operating in Sicily, Italy. The funder had no role in study design; the collection, analysis, and interpretation of data; the writing of the report; or in the decision to submit the article for publication. Researchers maintained independence from the funder and all authors had full access to all data (including statistical reports and tables) in the study and take responsibility for the integrity of the data and the accuracy of the data analysis.

## Data Availability

The patient data used in this study are available in an anonymised format on the Oxford University Research Archive (ORA). It can be accessed at the following DOI address: https://dx.doi.org/10.5287/ora-rjkzjgg4k.
As described in the data analysis software paragraph of the methods section, the code for this study was written in R version 4.4.1 using the tidyverse (v2.0.0), dplyr (v1.1.4), geepack (v1.3.13), lme4 (v1.1-35.5), and mice (v3.16.0) packages. All analysis code is available via GitHub at https://github.com/ethanp274/BOFE_trial.

## Acknowledgements

The authors gratefully acknowledge the support provided by Dr Gaetano Cardiel, President of SOFAD SRL; Dr Gioacchino Nicolosi, President of Federfarma Sicilia; the Sicilian region; and Dr Antonio Gaudioso, Secretary-General of CittadinanzAttiva. We also wish to acknowledge the pharmacists and patients who graciously supplied their time and energy in support of this trial.

## Notes

### Clinical Trial

ISRCTN 38734433

### Clinical Protocols

https://doi.org/10.1097/MD9.0000000000000158

### Author Declarations

Ethics approval for this study was obtained from relevant ethical review committees in both Italy and the United Kingdom. Approved 22/02/2021, Azienda Ospedaliero-Universitaria Policlinico “G. Rodolico – San Marco” Catania (via Santa Sofia 78 – 95123 Catania, Italy; +39 (0)95 3781855;) ref: 47/2021/PO Approved 29/03/2021, University of Central Lancashire (Preston Fylde Road, PR1 2HE, UK; +44 (0)1772 895583;) ref: HEALTH 0163

### Summary of Updates

Literature review shortened (Introduction); Strengths and limitations section shortened (Discussion); other revisions throughout for clarity, deduplication, and conciseness.

