## Supplementary Information for "Community pharmacist-led medicines use review for asthma and COPD: a randomised controlled trial of effectiveness and cost-effectiveness"

The patient data used in this study are available at https://dx.doi.org/10.5287/ora-rjkzjgg4k.

All code is available via GitHub at https://github.com/ethanp274/BOFE_trial.

### Supplementary Information S1: Additional Tables and Figures

**Supplementary Table 1.** Patient demographic and condition characteristics by condition and treatment arm.

| | Asthma patients (n=409) | | COPD patients (n=426) | | All patients (n=835) | |
|---|---|---|---|---|---|---|
| | **CRC-MUR** (n=266) | **Usual Care** (n=143) | **CRC-MUR** (n=294) | **Usual Care** (n=132) | **CRC-MUR** (n=560) | **Usual Care** (n=275) |
| Age: | | | | | | |
| Between 18 and 30 (%) | 9.40 | 9.09 | 0.00 | 0.00 | 4.46 | 4.73 |
| Between 31 and 40 (%) | 10.90 | 13.29 | 0.68 | 1.52 | 5.54 | 7.64 |
| Between 41 and 50 (%) | 16.17 | 16.78 | 2.72 | 1.52 | 9.11 | 9.45 |
| Between 51 and 60 (%) | 24.44 | 20.98 | 8.50 | 6.06 | 16.07 | 13.82 |
| Between 61 and 70 (%) | 19.92 | 20.98 | 28.57 | 36.36 | 24.46 | 28.36 |
| Between 71 and 80 (%) | 14.66 | 11.89 | 39.46 | 36.36 | 27.68 | 23.64 |
| Over 81 years old (%) | 4.51 | 6.99 | 20.07 | 18.18 | 12.68 | 12.36 |
| Male sex (%) | 39.85 | 45.45 | 69.05 | 71.21 | 55.18 | 57.82 |
| Height (m), mean (SD) | 1.65 (0.08) | 1.66 (0.10) | 1.66 (0.09) | 1.66 (0.08) | 1.66 (0.08) | 1.66 (0.09) |
| Weight (kg), mean (SD) | 72.41 (15.21) | 74.71 (16.30) | 75.25 (16.43) | 76.16 (19.18) | 73.90 (15.91) | 75.41 (17.72) |
| BMI, mean (SD) | 26.42 (5.09) | 27.24 (5.46) | 27.37 (5.46) | 27.55 (5.94) | 26.92 (5.30) | 27.39 (5.69) |
| Ethnicity: | | | | | | |
| Arabic / Middle Eastern (%) | 0.75 | 0.70 | 0.00 | 0.00 | 0.36 | 0.36 |
| Asian (%) | 0.00 | 0.70 | 0.00 | 0.00 | 0.00 | 0.36 |
| Black (%) | 0.00 | 0.00 | 0.68 | 0.00 | 0.36 | 0.00 |
| White / Caucasian (%) | 98.87 | 98.60 | 98.98 | 99.24 | 98.93 | 98.91 |
| Other | 0.38 | 0.00 | 0.34 | 0.76 | 0.36 | 0.36 |
| Location: | | | | | | |
| Agrigento (%) | 11.28 | 9.09 | 9.52 | 9.85 | 10.36 | 9.45 |
| Caltanissetta (%) | 4.14 | 5.59 | 7.14 | 7.58 | 5.71 | 6.55 |
| Catania (%) | 29.70 | 33.57 | 29.93 | 25.00 | 29.82 | 29.45 |
| Enna (%) | 5.64 | 4.90 | 6.12 | 6.82 | 5.89 | 5.82 |

| | | | | | | |
|---|---|---|---|---|---|---|
| Messina (%) | 17.67 | 13.99 | 14.97 | 17.42 | 16.25 | 15.64 |
| Palermo (%) | 13.16 | 13.99 | 13.27 | 15.15 | 13.21 | 14.55 |
| Ragusa (%) | 3.38 | 4.20 | 3.40 | 3.03 | 3.39 | 3.64 |
| Siracusa (%) | 11.28 | 10.49 | 12.24 | 11.36 | 11.79 | 10.91 |
| Trapani (%) | 3.76 | 4.20 | 3.40 | 3.79 | 3.57 | 4.00 |
| Recruitment Source: | | | | | | |
| Pharmacist (%) | 89.85 | 88.81 | 89.46 | 91.67 | 89.64 | 90.18 |
| GP (%) | 10.15 | 11.19 | 10.20 | 8.33 | 10.18 | 9.82 |
| Consultant/Specialist (%) | 0.00 | 0.00 | 0.34 | 0.00 | 0.18 | 0.00 |
| Live alone (%) | 15.04 | 13.99 | 19.39 | 15.91 | 17.32 | 14.91 |
| Employed (%) | 54.14 | 53.85 | 83.33 | 88.64 | 69.46 | 70.55 |
| Highest education attained: | | | | | | |
| No school (%) | 0.36 | 1.40 | 1.36 | 0.76 | 0.89 | 1.09 |
| Primary school (%) | 11.65 | 12.59 | 30.61 | 35.61 | 21.61 | 23.64 |
| Secondary school (%) | 64.66 | 65.73 | 55.44 | 54.55 | 59.82 | 60.36 |
| University (%) | 23.31 | 20.28 | 12.59 | 9.09 | 17.68 | 14.91 |
| Pneumonia vaccine in previous 12mo (%) | 40.98 | 45.45 | 70.07 | 74.24 | 56.25 | 59.27 |
| COVID vaccine in previous 12mo (%) | 17.29 | 13.99 | 32.31 | 33.33 | 25.18 | 23.27 |
| Heart disease (%) | 9.40 | 12.59 | 20.75 | 23.48 | 15.38 | 17.82 |
| Other chronic disease (%) | 29.32 | 31.47 | 57.14 | 65.15 | 43.93 | 47.64 |
| Smoking status: | | | | | | |
| Yes (%) | 17.67 | 18.18 | 21.77 | 19.70 | 19.82 | 18.91 |
| Yes - occasionally (%) | 6.39 | 10.49 | 4.08 | 6.06 | 5.18 | 8.36 |
| Not anymore (%) | 27.44 | 25.17 | 55.78 | 57.58 | 42.32 | 40.73 |
| Never (%) | 48.50 | 46.15 | 18.37 | 16.67 | 32.68 | 32.00 |
| Number of medications, mean (SD) | 5.03 (3.34) | 4.71 (3.29) | 6.59 (3.40) | 7.23 (3.91) | 5.85 (3.46) | 5.92 (3.81) |
| Medication issues: | | | | | | |
| No problems (%) | 84.96* | 93.01* | 82.99 | 84.09 | 83.93 | 88.73 |
| Some problems (%) | 15.04* | 6.29* | 15.99 | 14.39 | 15.54 | 10.18 |
| Lots of problems (%) | 0.00* | 0.70* | 1.02 | 1.52 | 0.54 | 1.09 |
| Medication understanding: | | | | | | |
| Full understanding (%) | 86.47 | 85.31 | 84.35 | 82.58 | 85.36 | 84.00 |
| Partial understanding (%) | 13.16 | 13.29 | 13.61 | 14.39 | 13.39 | 13.82 |
| Does not understand (%) | 0.38 | 1.40 | 2.04 | 3.03 | 1.25 | 2.18 |
| ACT Score at baseline, mean (SD) | 18.48 (4.30) | 19.20 (4.44) | NA | NA | NA | NA |
| CCQ Score at baseline, mean (SD) | NA | NA | 1.99* (1.16) | 2.25* (1.29) | NA | NA |
| Quality of life index, mean (SD) | 0.856 (0.172) | 0.865 (0.178) | 0.770 (0.271) | 0.732 (0.308) | 0.811 (0.233) | 0.801 (0.257) |

**Significant difference across treatment arms (p<0.05)*

**Supplementary Table 2.** Health and quality of life outcomes among patients with asthma by treatment arm.

| | CRC-MUR | | | | | | Usual Care | | | | | |
|---|---|---|---|---|---|---|---|---|---|---|---|---|
| | T0 (n=266) | T3 (n=260) | T6 (n=254) | T9 (n=257) | T12 (n=251) | T12–T0 | T0 (n=143) | T3 (n=140) | T6 (n=141) | T9 (n=141) | T12 (n=140) | T12–T0 |
| Asthma Controlled (%) | 46.24* | 53.46 | 57.87 | 54.09 | 53.78 | +7.54 | 58.74* | 50.71 | 58.16 | 50.35 | 55.71 | -3.03 |
| ACT score, mean (SD) | 18.48 (4.30) | 19.30 (4.42) | 19.46 (4.50) | 19.52 (4.34) | 19.36 (4.54) | +0.88 (4.82) | 19.20 (4.44) | 19.00 (4.62) | 19.44 (4.86) | 18.79 (4.70) | 19.64 (4.38) | +0.33 (4.37) |
| EQ5D index, mean (SD) | 0.856 (0.172) | 0.878 (0.178) | 0.868* (0.205) | 0.893 (0.166) | 0.884 (0.178) | +0.024 (0.170) | 0.865 (0.178) | 0.884 (0.175) | 0.907* (0.161) | 0.895 (0.169) | 0.896 (0.156) | +0.027 (0.162) |

**Significant difference across treatment arms (p<0.05)*

**Supplementary Table 3.** Health and quality of life outcomes among patients with COPD by treatment arm.

| | CRC-MUR | | | | | | Usual Care | | | | | |
|---|---|---|---|---|---|---|---|---|---|---|---|---|
| | T0 (n=294) | T3 (n=284) | T6 (n=274) | T9 (n=267) | T12 (n=254) | T12–T0 | T0 (n=132) | T3 (n=129) | T6 (n=127) | T9 (n=127) | T12 (n=123) | T12–T0 |
| COPD Controlled (%) | 55.44 | 59.51 | 56.93* | 59.55 | 62.20* | +6.76 | 47.73 | 49.61 | 45.67* | 49.61 | 49.59* | +1.86 |
| CCQ score, mean (SD) | 1.99* (1.16) | 1.90* (1.21) | 1.93* (1.20) | 1.82* (1.21) | 1.78* (1.22) | -0.15 (1.15) | 2.25* (1.29) | 2.16* (1.27) | 2.31* (1.44) | 2.17* (1.47) | 2.17* (1.38) | -0.08 (1.03) |
| EQ5D index, mean (SD) | 0.770 (0.271) | 0.789 (0.265) | 0.799* (0.268) | 0.793* (0.272) | 0.810* (0.270) | +0.020 (0.239) | 0.732 (0.308) | 0.735 (0.331) | 0.734* (0.315) | 0.720* (0.352) | 0.718* (0.342) | -0.010 (0.265) |

**Significant difference across treatment arms (p<0.05)*

**Supplementary Table 4.** Health and social care resource utilization across time points by treatment arm.

| | CRC-MUR | | | | Usual Care | | | |
|---|---|---|---|---|---|---|---|---|
| (all variables capture prior 6 months) | T0 (n=560) | T6 (n=528) | T12 (n=505) | T12-T0 | T0 (n=275) | T6 (n=268) | T12 (n=263) | T12-T0 |
| GP Visits, mean (SD) | 3.35 (3.97) | 2.69 (3.20) | 2.58 (3.58) | -0.63 (4.44) | 3.29 (5.06) | 3.30 (6.33) | 2.84 (3.87) | -0.41 (3.92) |
| Nurse Visits, mean (SD) | 0.13 (0.85) | 0.12 (0.76) | 0.10 (0.73) | 0.00 (0.94) | 0.08 (0.50) | 0.27 (1.55) | 0.18 (0.86) | +0.12 (0.87) |
| Therapist Visits, mean (SD) | 1.29* (6.18) | 0.42 (2.06) | 0.55 (2.67) | -0.75 (6.38) | 0.57* (2.23) | 0.29 (1.63) | 0.28 (1.48) | -0.27 (2.25) |
| A&E Visits, mean (SD) | 0.19 (0.53) | 0.11 (0.43) | 0.11 (0.47) | -0.08 (0.68) | 0.16 (0.53) | 0.13 (0.44) | 0.16 (0.59) | +0.01 (0.64) |
| Hospital (Outpatient) Visits, mean (SD) | 0.83 (1.67) | 0.50 (1.17) | 0.48 (0.98) | -0.34 (1.69) | 0.68 (1.07) | 0.60 (1.39) | 0.55 (1.19) | -0.12 (1.44) |
| Hospital (Inpatient) Visits, mean (SD) | 0.32 (1.06) | 0.18 (0.60) | 0.21 (0.91) | -0.11 (1.35) | 0.27 (1.10) | 0.21 (0.75) | 0.22 (0.77) | -0.04 (1.24) |
| Hospital (Inpatient) Days, mean (SD) | 0.90 (3.69) | 0.58 (3.32) | 0.67 (4.46) | -0.24 (5.61) | 1.12 (5.42) | 0.57 (2.92) | 0.87 (3.84) | -0.28 (6.07) |
| Social Worker Visits, mean (SD) | 0.08 (0.63) | 0.09 (1.09) | 0.05 (0.43) | -0.04 (0.79) | 0.21 (1.69) | 0.28 (3.02) | 0.06 (0.65) | -0.13 (1.18) |
| Adult Day Centre Visits (per week), mean (SD) | 0.07 (1.52) | 0.02 (0.18) | 0.02 (0.32) | -0.06 (1.64) | 0.01 (0.10) | 0.03 (0.26) | 0.02 (0.16) | +0.01 (0.20) |

**Significant difference across treatment arms (p<0.05)*

**Supplementary Table 5.** Costs incurred by the national health service over the 12-month study period by treatment arm.

| | **CRC-MUR** (n = 473) | **Usual Care** (n = 254) | **p-value**[†] |
|---|---|---|---|
| Outpatient Costs (€), mean (SD) | 328 (774) | 474 (2960) | 0.675 |
| Laboratory Costs (€), mean (SD) | 348 (720.) | 337 (658) | 0.686 |
| Medication Costs (€), mean (SD) | 961 (930.) | 1050 (1220) | 0.748 |
| Medication Delivery Costs (€), mean (SD) | 392 (2460) | 475 (1810) | 0.095 |
| Hospital (Inpatient) Costs (€), mean (SD) | 882 (3040) | 1170 (4250) | 0.939 |
| Total Costs (€), mean (SD) | 4430 (7220) | 5560 (10980) | 0.751 |

[†]*Testing difference between groups at 12mo using Wilcoxon rank-sum (Mann-Whitney) test*

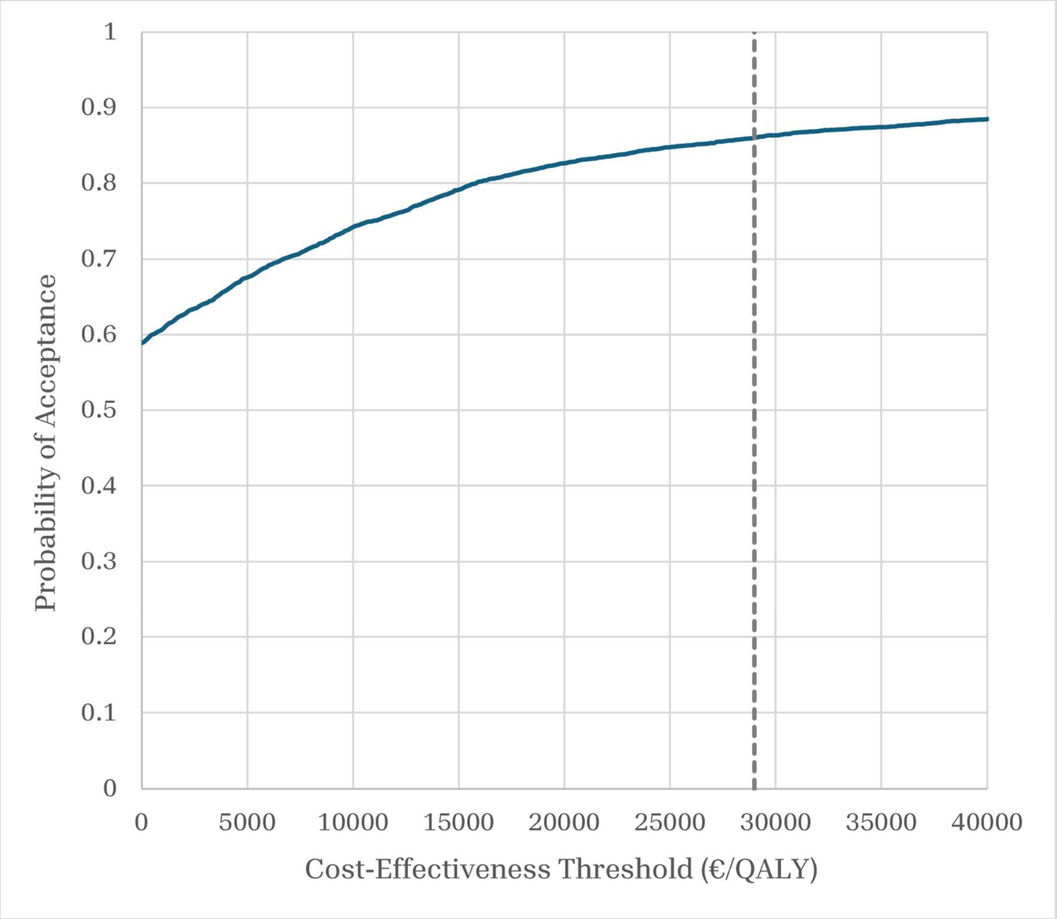


**Supplementary Figure 1**. Cost-effectiveness acceptability curve for CRC-MUR. Vertical dashed line represents a cost-effectiveness threshold of 29,000 EUR/QALY (approximately 25,000 GBP/QALY). Probability of acceptance at displayed threshold is 86.1%.

### Supplementary Information S2: Robustness and Sensitivity Analyses

Overview

To evaluate the robustness of the findings, we conducted a series of post hoc sensitivity analyses examining alternative assumptions relating to missing data handling, model specification, clustering, cost-effectiveness estimation, intervention costing, and quality-of-life valuation. Sensitivity analyses were intended to assess the stability of the findings under defensible alternative assumptions and were not used to select preferred models.

For effectiveness outcomes, sensitivity analyses were grouped into two broad categories: (1) alternative approaches to handling missing outcome data and specifying MICE predictor matrices, and (2) alternative outcome-model specifications examining longitudinal structure, covariate adjustment, and clustering assumptions. For economic outcomes, sensitivity analyses examined alternative missing-data assumptions, health utility valuation methods, and intervention-cost assumptions.

Collectively, these sensitivity analyses indicate that the primary findings were robust to alternative assumptions regarding missing data, model specification, clustering structure, intervention costs, and health utility valuation. While the magnitude and precision of the estimated effects varied modestly across specifications, the direction of both the effectiveness and cost-effectiveness findings remained consistent throughout.

S2.1: Sensitivity to Missing-Data Handling and MICE Predictor-Matrix Specification

The primary effectiveness analysis used multivariate imputation by chained equations (MICE) with a rule-based, time-aware predictor matrix that incorporated demographic characteristics, disease-control history, quality-of-life measures, and other clinically relevant predictors while prohibiting future-timepoint information. Imputation in this way allows us to follow an intention-to-treat (ITT) approach, as specified in the protocol (1). To assess whether the intervention effect depended on assumptions regarding missing-data handling or predictor selection, we compared the primary MICE specification against several alternative approaches. These included complete-case analysis (excluding participants with incomplete follow-up data), naïve within-arm mean or mode imputation, a time-sanitised empirical predictor matrix generated using the “quickpred()” algorithm (2), and more restrictive MICE specifications that removed utility-related auxiliary variables, removed same-visit auxiliary variables, or restricted predictors to historical disease-control information only. All imputation approaches were performed separately within treatment arms to avoid treatment-effect leakage and were restricted to baseline or prior-timepoint information when imputing later outcomes. These analyses assess whether conclusions depend on assumptions regarding missing-data mechanisms, auxiliary-variable inclusion, or predictor-matrix complexity. Results are presented in Supplementary Table 6.

**Supplementary Table 6.** CRC-MUR effectiveness estimates for disease control at 12-months under alternative missing-data and MICE predictor-matrix specifications.

| Missing-data approach | Predictor links | Adjusted Odds Ratio | P-value |
|---|---|---|---|
| Complete case analysis | NA | 1.462 [1.039, 2.058] | 0.029 |
| Simple within-arm imputation (mean/mode) | NA | 1.564 [1.128, 2.168] | 0.007 |
| Time-sanitised "quickpred" MICE | 63 | 1.403 [0.999, 1.970] | 0.051 |
| History-only disease control MICE | 117 | 1.443 [1.029, 2.024] | 0.033 |
| No same-visit variables MICE | 130 | 1.396 [0.999, 1.951] | 0.051 |
| No utility-index variables MICE | 121 | 1.419 [1.016, 1.982] | 0.040 |
| Main analysis MICE | 152 | 1.414 [1.010, 1.979] | 0.044 |

### S2.2: Sensitivity to Outcome-Model Specification, Longitudinal Structure, and Pharmacy-Level Clustering

The primary effectiveness analysis used an adjusted marginal logistic generalised estimating equation (GEE) model with disease control at follow-up as the outcome and baseline disease control, age, sex, treatment group, time, and treatment-by-time interaction as predictors. This approach followed the pre-specified analytic plan described in the study protocol (1). Sensitivity analyses were conducted to examine whether the estimated intervention effect depended on model structure, degree of adjustment, or clustering assumptions. Alternative specifications included unadjusted GEE models, cross-sectional logistic regression models restricted to the 12-month endpoint, mixed-effects logistic regression models with patient-level random intercepts, and mixed-effects models incorporating both patient-level and pharmacy-clustered random intercepts. A maximally adjusted GEE specification was also estimated, extending the primary model to include additional baseline covariates available in the trial dataset, including BMI, smoking status, condition type, and ischaemic heart disease. These analyses evaluate whether conclusions are sensitive to assumptions regarding repeated-measures correlation, subject-specific versus population-average estimands, baseline covariate adjustment, or clustering by pharmacy. Results are presented in Supplementary Table 7.

Differences between the GEE and mixed-effects estimates are expected because the models target different estimands. The GEE model estimates a population-average intervention effect, whereas the mixed-effects models estimate subject-specific effects conditional on random effects. In logistic regression, subject-specific odds ratios are typically further from the null than marginal odds ratios because of the non-collapsibility of the odds ratio (3,4).

**Supplementary Table 7.** CRC-MUR effectiveness estimates for disease control at 12-months using different model structures and specifications.

| Model | Odds Ratio | P-value |
|---|---|---|
| Unadjusted GEE | 1.229 [0.913, 1.654] | 0.173 |
| Unadjusted GLM (non-longitudinal) | 1.229 [0.913, 1.655] | 0.174 |
| Adjusted GEE (used in main analysis) | 1.414 [1.010, 1.979] | 0.044 |
| Maximally adjusted GEE | 1.386 [0.987, 1.948] | 0.060 |
| Adjusted GLM (non-longitudinal) | 1.392 [0.998, 1.941] | 0.052 |
| Adjusted GLMM (patient random effects) | 1.734 [1.021, 2.945] | 0.042 |
| Adjusted clustered GLMM (patient and pharmacy random effects) | 1.687 [1.016, 2.802] | 0.043 |

### S2.3: Sensitivity to Missing-Data Handling in the Cost-Effectiveness Analysis

The primary cost-effectiveness analysis used a dedicated MICE specification designed for economic evaluation, incorporating demographic characteristics, disease-control outcomes, EQ-5D responses, and health-system cost variables. This approach allowed us to maintain the ITT approach used for the main effectiveness analysis. To assess whether economic conclusions depended on missing-data assumptions, we compared the primary analysis against complete-case analysis and naïve within-arm mean or mode imputation. Complete-case analysis excludes all participants with missing cost or quality-of-life data, whereas simple imputation replaces missing values using within-arm averages or modal values. These approaches represent alternative assumptions regarding the missing-data process and provide a stress test for the robustness of the economic findings. Results are presented in Supplementary Table 8.

**Supplementary Table 8.** Cost-effectiveness results using different approaches for addressing missing data.

| Missing-data approach | Incremental Cost (€) | Incremental QALYs | Probability Cost-Effective |
|---|---|---|---|
| MICE (used in main analysis) | -116.75 | 0.0224 | 0.861 |
| Simple imputation (mean/mode) | -123.27 | 0.0282 | 0.903 |
| Complete case analysis | -469.93 | 0.0322 | 0.971 |

S2.4: Sensitivity to Health Utility Valuation (EQ-5D Tariff Selection)

The primary cost-effectiveness analysis calculated QALYs using the Italian EQ-5D-5L value set (5), which reflects health-state preferences elicited from the Italian population. To assess whether economic conclusions depended on the choice of health utility tariff, QALYs were recalculated using the United Kingdom EQ-5D-5L value set (6). Because tariff selection affects the valuation of health states but not healthcare costs, this analysis specifically evaluates the sensitivity of QALY estimates and cost-effectiveness conclusions to alternative population preference weights. Results are presented in Supplementary Table 9.

**Supplementary Table 9.** Incremental QALYs and cost-effectiveness probabilities under varied quality of life tariff sets.

| EQ-5D-5L tariff set | Incremental QALYs | Probability Cost-Effective |
|---|---|---|
| Italy (used in main analysis) | 0.022 [-0.020, 0.065] | 0.861 |
| England | 0.019 [-0.020, 0.059] | 0.835 |

S2.5: Sensitivity to Intervention-Cost Assumptions

The primary economic analysis assumed an intervention delivery cost of €40 per consultation, consistent with the costing framework used in the earlier Italian Medicines Use Review (I-MUR) trial (7). Because the true reimbursement level for pharmacist-led medicines use review services may vary across healthcare systems and implementation models, we evaluated cost-effectiveness across a range of alternative consultation costs from €40 to €200 per consultation. These analyses assess whether the economic conclusions remain robust under more conservative assumptions regarding intervention delivery costs. Results are presented in Supplementary Table 10.

**Supplementary Table 10.** Probability of CRC-MUR cost-effectiveness under varied consultation costs.

| Cost per consultation (€) | Incremental Cost (€) | Probability Cost-Effective |
|---|---|---|
| 40 (used in main analysis) | -116.75 | 0.861 |
| 80 | -36.75 | 0.836 |
| 120 | 43.25 | 0.807 |
| 160 | 123.25 | 0.774 |
| 200 | 203.25 | 0.736 |

### Supplementary Information S3: CONSORT Checklist for Reporting Clinical Trials

CONSORT 2025 checklist of information to include when reporting a randomised trial (8).

| Section / Topic | No | CONSORT 2025 checklist item description | Reported on page no. |
|---|---|---|---|
| **Title and abstract** | | | |
| Title and structured abstract | 1a | Identification as a randomised trial | 1 |
| | 1b | Structured summary of the trial design, methods, results, and conclusions | 2 |
| **Open science** | | | |
| Trial registration | 2 | Name of trial registry, identifying number (with URL) and date of registration | 2, 15 |
| Protocol and statistical analysis plan | 3 | Where the trial protocol and statistical analysis plan can be accessed | 4, 15 |
| Data sharing | 4 | Where and how the individual de-identified participant data (including data dictionary), statistical code and any other materials can be accessed | 15 |
| Funding and conflicts of interest | 5a | Sources of funding and other support (e.g., supply of drugs), and role of funders in the design, conduct, analysis and reporting of the trial | 16 |
| | 5b | Financial and other conflicts of interest of the manuscript authors | 15 |
| **Introduction** | | | |
| Background and rationale | 6 | Scientific background and rationale | 3-4 |
| Objectives | 7 | Specific objectives related to benefits and harms | 4, 5-6 |
| **Methods** | | | |
| Patient and public involvement | 8 | Details of patient or public involvement in the design, conduct and reporting of the trial | 6 |

| | | | |
|---|---|---|---|
| Trial design | 9 | Description of trial design including type of trial (e.g., parallel group, crossover), allocation ratio, and framework (e.g., superiority, equivalence, non-inferiority, exploratory) | 4-5 |
| Changes to trial protocol | 10 | Important changes to the trial after it commenced including any outcomes or analyses that were not prespecified, with reason | 7-8 & Supplementary Information (post-hoc sensitivity analyses) |
| Trial setting | 11 | Settings (e.g., community, hospital) and locations (e.g., countries, sites) where the trial was conducted | 4 |
| Eligibility criteria | 12a | Eligibility criteria for participants | 4 |
| | 12b | If applicable, eligibility criteria for sites and for individuals delivering the interventions (e.g., surgeons, physiotherapists) | 4 |
| Intervention and comparator | 13 | Intervention and comparator with sufficient details to allow replication. If relevant, where additional materials describing the intervention and comparator (e.g., intervention manual) can be accessed | 5-6 |
| Outcomes | 14 | Pre-specified primary and secondary outcomes, including the specific measurement variable (e.g., systolic blood pressure), analysis metric (e.g., change from baseline, final value, time to event), method of aggregation (e.g., median, proportion), and time point for each outcome | 7-8 |
| Harms | 15 | How harms were defined and assessed (e.g., systematically, non-systematically) | 5 |
| Sample size | 16a | How sample size was determined, including all assumptions supporting the sample size calculation | 4-5 |
| | 16b | Explanation of any interim analyses and stopping guidelines | NA |
| Randomisation: | | | |
| Sequence generation | 17a | Who generated the random allocation sequence and the method used | 5 |
| | 17b | Type of randomisation and details of any restriction (e.g., stratification, blocking and block size) | 5 |

| | | | |
|---|---|---|---|
| Allocation concealment mechanism | 18 | Mechanism used to implement the random allocation sequence (e.g., central computer/telephone; sequentially numbered, opaque, sealed containers), describing any steps to conceal the sequence until interventions were assigned | 5 |
| Implementation | 19 | Whether the personnel who enrolled and those who assigned participants to the interventions had access to the random allocation sequence | 5 |
| Blinding | 20a | Who was blinded after assignment to interventions (e.g., participants, care providers, outcome assessors, data analysts) | 5 |
| | 20b | If blinded, how blinding was achieved and description of the similarity of interventions | NA |
| Statistical methods | 21a | Statistical methods used to compare groups for primary and secondary outcomes, including harms | 7-8 |
| | 21b | Definition of who is included in each analysis (e.g., all randomised participants), and in which group | 7-8 |
| | 21c | How missing data were handled in the analysis | 7 |
| | 21d | Methods for any additional analyses (e.g., subgroup and sensitivity analyses), distinguishing prespecified from post-hoc | 7-8 & supplementary information |
| **Results** | | | |
| Participant flow, including flow diagram | 22a | For each group, the numbers of participants who were randomly assigned, received intended intervention, and were analysed for the primary outcome | 8, Figure 1 |
| | 22b | For each group, losses and exclusions after randomisation, together with reasons | Figure 1 |
| Recruitment | 23a | Dates defining the periods of recruitment and follow-up for outcomes of benefits and harms | 8 |
| | 23b | If relevant, why the trial ended or was stopped | NA |
| Intervention and comparator delivery | 24a | Intervention and comparator as they were actually administered (e.g., where appropriate, who delivered the intervention/comparator, how participants adhered, whether they were delivered as intended [fidelity]) | NA (fidelity monitoring not undertaken) |

| | | | |
|---|---|---|---|
| | 24b | Concomitant care received during the trial for each group | Supplementary Table 4 |
| Baseline data | 25 | A table showing baseline demographic and clinical characteristics for each group | Table 1 & Supplementary Table 1 |
| Numbers analysed, outcomes and estimation | 26 | For each primary and secondary outcome, by group: <br>• the number of participants included in the analysis <br>• the number of participants with available data at the outcome time point <br>• result for each group, and the estimated effect size and its precision (such as 95% confidence interval) <br>• for binary outcomes, presentation of both absolute and relative effect size | 8-10, Tables 2-3 |
| Harms | 27 | All harms or unintended events in each group | NA (None) |
| Ancillary analyses | 28 | Any other analyses performed, including subgroup and sensitivity analyses, distinguishing pre-specified from post-hoc | 9-10 & Supplementary information |
| **Discussion** | | | |
| Interpretation | 29 | Interpretation consistent with results, balancing benefits and harms, and considering other relevant evidence | 10, 12-14 |
| Limitations | 30 | Trial limitations, addressing sources of potential bias, imprecision, generalisability, and, if relevant, multiplicity of analyses | 10-12 |

© 2025 Hopewell et al. This is an Open Access article distributed under the terms of the Creative Commons Attribution License (https://creativecommons.org/licenses/by/4.0/), which permits unrestricted use, distribution, and reproduction in any medium, provided the original work is properly cited.

### Supplementary Information S4: CHEERS Checklist for Reporting Health Economic Evaluations

CHEERS 2022 checklist of information to include when reporting a health economic evaluation (9).

| Topic | No. | Item | Location where item is reported |
|---|---|---|---|
| **Title** | | | |
| | 1 | Identify the study as an economic evaluation and specify the interventions being compared. | Title (p1) |
| **Abstract** | | | |
| | 2 | Provide a structured summary that highlights context, key methods, results, and alternative analyses. | Abstract (p2) |
| **Introduction** | | | |
| **Background and objectives** | 3 | Give the context for the study, the study question, and its practical relevance for decision making in policy or practice. | Introduction (p3-4) |
| **Methods** | | | |
| **Health economic analysis plan** | 4 | Indicate whether a health economic analysis plan was developed and where available. | Methods (p4) |
| **Study population** | 5 | Describe characteristics of the study population (such as age range, demographics, socioeconomic, or clinical characteristics). | Methods (p4-5) |
| **Setting and location** | 6 | Provide relevant contextual information that may influence findings. | Methods (p4-5) |
| **Comparators** | 7 | Describe the interventions or strategies being compared and why chosen. | Methods (p5-6) |
| **Perspective** | 8 | State the perspective(s) adopted by the study and why chosen. | Methods (p6) |
| **Time horizon** | 9 | State the time horizon for the study and why appropriate. | Methods (p6) |
| **Discount rate** | 10 | Report the discount rate(s) and reason chosen. | Methods (p6) |

| Topic | No. | Item | Location where item is reported |
|---|---|---|---|
| **Selection of outcomes** | 11 | Describe what outcomes were used as the measure(s) of benefit(s) and harm(s). | Methods (p7) |
| **Measurement of outcomes** | 12 | Describe how outcomes used to capture benefit(s) and harm(s) were measured. | Methods (p7) |
| **Valuation of outcomes** | 13 | Describe the population and methods used to measure and value outcomes. | Methods (p6-8) |
| **Measurement and valuation of resources and costs** | 14 | Describe how costs were valued. | Methods (p8) |
| **Currency, price date, and conversion** | 15 | Report the dates of the estimated resource quantities and unit costs, plus the currency and year of conversion. | Methods (p6) |
| **Rationale and description of model** | 16 | If modelling is used, describe in detail and why used. Report if the model is publicly available and where it can be accessed. | Methods (p7-8), Endmatter (p15) |
| **Analytics and assumptions** | 17 | Describe any methods for analysing or statistically transforming data, any extrapolation methods, and approaches for validating any model used. | Methods (p7-8) |
| **Characterising heterogeneity** | 18 | Describe any methods used for estimating how the results of the study vary for subgroups. | Not undertaken/applicable |
| **Characterising distributional effects** | 19 | Describe how impacts are distributed across different individuals or adjustments made to reflect priority populations. | Not undertaken/applicable |
| **Characterising uncertainty** | 20 | Describe methods to characterise any sources of uncertainty in the analysis. | Methods (p8) |
| **Approach to engagement with patients and others affected by the study** | 21 | Describe any approaches to engage patients or service recipients, the general public, communities, or stakeholders (such as clinicians or payers) in the design of the study. | Methods (p6) |
| **Results** | | | |
| **Study parameters** | 22 | Report all analytic inputs (such as values, ranges, references) including uncertainty or distributional assumptions. | Not applicable |
| **Summary of main results** | 23 | Report the mean values for the main categories of costs and outcomes of interest and summarise them in the most appropriate overall measure. | Results (p9), Supplementary Information |

| Topic | No. | Item | Location where item is reported |
|---|---|---|---|
| **Effect of uncertainty** | 24 | Describe how uncertainty about analytic judgments, inputs, or projections affect findings. Report the effect of choice of discount rate and time horizon, if applicable. | Results (p9-10) |
| **Effect of engagement with patients and others affected by the study** | 25 | Report on any difference patient/service recipient, general public, community, or stakeholder involvement made to the approach or findings of the study | Not undertaken/applicable |
| **Discussion** | | | |
| **Study findings, limitations, generalisability, and current knowledge** | 26 | Report key findings, limitations, ethical or equity considerations not captured, and how these could affect patients, policy, or practice. | Discussion (p10-14) |
| **Other relevant information** | | | |
| **Source of funding** | 27 | Describe how the study was funded and any role of the funder in the identification, design, conduct, and reporting of the analysis | Endmatter (p16) |
| **Conflicts of interest** | 28 | Report authors conflicts of interest according to journal or International Committee of Medical Journal Editors requirements. | Endmatter (p15) |
